# SARS-CoV-2 RNAaemia predicts clinical deterioration and extrapulmonary complications from COVID-19

**DOI:** 10.1101/2020.12.19.20248561

**Authors:** Nikhil Ram-Mohan, David Kim, Elizabeth J Zudock, Marjan M Hashemi, Kristel C Tjandra, Angela J Rogers, Catherine A Blish, Kari C. Nadeau, Jennifer A Newberry, James V Quinn, Ruth O’Hara, Euan Ashley, Hien Nguyen, Lingxia Jiang, Paul Hung, the Stanford COVID-19 Biobank Study Group, Andra L Blomkalns, Samuel Yang

**Affiliations:** Department of Emergency Medicine, Stanford University School of Medicine, Palo Alto CA 94305; Department of Medicine - Pulmonary, Allergy & Critical Care Medicine, Stanford University School of Medicine, Palo Alto CA 94305; Department of Medicine/Infectious Diseases, Stanford University School of Medicine, Palo Alto CA 94305; Department of Psychiatry and Behavioral Sciences, Stanford University School of Medicine, Palo Alto CA 94305; Department of Medicine – Cardiovascular Medicine, Stanford University School of Medicine, Palo Alto CA 94305; COMBiNATi Inc., 2450 Embarcadero Way, Palo Alto CA 94303

## Abstract

**Background:** The determinants of COVID-19 disease severity and extrapulmonary complications (EPCs) are poorly understood. We characterise the relationships between SARS-CoV-2 RNAaemia and disease severity, clinical deterioration, and specific EPCs.

**Methods:** We used quantitative (qPCR) and digital (dPCR) PCR to quantify SARS-CoV-2 RNA from nasopharyngeal swabs and plasma in 191 patients presenting to the Emergency Department (ED) with COVID-19. We recorded patient symptoms, laboratory markers, and clinical outcomes, with a focus on oxygen requirements over time. We collected longitudinal plasma samples from a subset of patients. We characterised the role of RNAaemia in predicting clinical severity and EPCs using elastic net regression.

**Findings:** 23·0% (44/191) of SARS-CoV-2 positive patients had viral RNA detected in plasma by dPCR, compared to 1·4% (2/147) by qPCR. Most patients with serial measurements had undetectable RNAaemia 10 days after onset of symptoms, but took 16 days to reach maximum severity, and 33 days for symptoms to resolve. Initially RNAaemic patients were more likely to manifest severe disease (OR 6·72 [95% CI, 2·45 – 19·79]), worsening of disease severity (OR 2·43 [95% CI, 1·07 - 5·38]), and EPCs (OR 2·81 [95% CI, 1·26 – 6·36]). RNA load correlated with maximum severity (*r* = 0·47 [95% CI, 0·20 - 0·67]).

**Interpretation:** dPCR is more sensitive than qPCR for the detection of SARS-CoV-2 RNAaemia, which is a robust predictor of eventual COVID-19 severity and oxygen requirements, as well as EPCs. Since many COVID-19 therapies are initiated on the basis of oxygen requirements, RNAaemia on presentation might serve to direct early initiation of appropriate therapies for the patients most likely to deteriorate.

**Funding:** NIH/NIAID (Grants R01A153133, R01AI137272, and 3U19AI057229 – 17W1 COVID SUPP #2) and a donation from Eva Grove.

**Research in context:** *Evidence before this study:* The varied clinical manifestations of COVID-19 have directed attention to the distribution of SARS-CoV-2 in the body. Although most concentrated and tested for in the nasopharynx, SARS-CoV-2 RNA has been found in blood, stool, and numerous tissues, raising questions about dissemination of viral RNA throughout the body, and the role of this process in disease severity and extrapulmonary complications. Recent studies have detected low levels of SARS-CoV-2 RNA in blood using either quantitative reverse transcriptase real-time PCR (qPCR) or droplet digital PCR (dPCR), and have associated RNAaemia with disease severity and biomarkers of dysregulated immune response.

*Added value of this study:* We quantified SARS-CoV-2 RNA in the nasopharynx and plasma of patients presenting to the Emergency Department with COVID-19, and found an array-based dPCR platform to be markedly more sensitive than qPCR for detection of SARS-CoV-2 RNA, with a simplified workflow well-suited to clinical adoption. We collected serial plasma samples during patients’ course of illness, and showed that SARS-CoV-2 RNAaemia peaks early, while clinical condition often continues to worsen. Our findings confirm the association between RNAaemia and disease severity, and additionally demonstrate a role for RNAaemia in predicting future deterioration and specific extrapulmonary complications.

*Implications of all the available evidence:* Variation in SARS-CoV-2 RNAaemia may help explain disparities in disease severity and extrapulmonary complications from COVID-19. Testing for RNAaemia with dPCR early in the course of illness may help guide patient triage and management.

## Introduction

As of December 2020, SARS-CoV-2 has caused over 70 million infections and 1·6 million deaths.^1^ The variability of patient responses to COVID-19 makes it difficult for frontline clinicians to identify and appropriately triage patients most at risk for clinical deterioration. While COVID-19 is often manifest as a viral pneumonia, multi-organ involvement can produce more severe and recalcitrant disease.^2,3^ SARS-CoV-2, typically isolated from nasopharyngeal (NP) samples, has been detected in lower titers in whole blood, serum, plasma, and stool.^4–10^ Histopathological surveys have identified the virus in myocardial, renal, gastrointestinal, and neurological tissues.^11–14^ Is haematogenous spread of the virus or viral components associated with extrapulmonary complications (EPCs)? The clinical and pathophysiological significance of SARS-CoV-2 RNA in blood remains poorly understood.

SARS-CoV-2 RNAaemia, detected with quantitative reverse transcriptase real-time PCR (qPCR), has been correlated with severity of COVID-19. However, reported rates of RNA detection range from 0% – 41% in serum and 13% – 33% in plasma.^4,6,7,10,15–17^ Comparing across studies is challenging, due to discrepancies in assay protocols and sensitivities, collections timings, and patient populations. Conventional qPCR also lacks the sensitivity and precision to reliably detect and measure low viral loads.^18^ Digital PCR (dPCR) offers improved sensitivity, precision, and reproducibility over qPCR, with absolute quantification of viral RNA without standard curves. Given its tolerance to inhibitors, dPCR is particularly suited to detecting dilute targets in blood.^19^ Two cross-sectional studies using droplet-based dPCR reported higher rates of RNAaemia (42·4% and 74·1%) than did the qPCR studies above, and associated the presence and the level of RNAaemia with clinical severity.^8,9^ Bermejo-Martin *et al* also observed correlation between RNAaemia and biomarkers of dysregulated host responses.^8^

In this prospective, longitudinal, observational study of COVID-19 patients presenting to the Emergency Department (ED), we characterised relationships between SARS-CoV-2 RNAaemia and overall severity, clinical deterioration, and specific EPCs. We used an array-based dPCR platform to maximise reliability and replicability, and to simplify potential clinical adoption of RNAaemia testing.

## Materials and Methods

### COVID-19 patients and specimen collection

We collected peripheral blood +/- NP swabs from patients prospectively enrolled in the IRB-approved (eP-55650) Stanford University ED COVID-19 Biobank beginning in April 2020, after written informed consent from patients or their surrogates. Eligibility criteria were age ≥18 years and presentation to the Stanford Hospital ED with a positive screening SARS-CoV-2 NP swab, analysed by RT-PCR as part of routine ED care. We repeated blood draws on one or more of days three, seven and 30 if the patient remained hospitalised. We asked discharged participants to return for repeat blood draws on days seven and 30 from enrollment. We collected blood in ethylenediaminetetraacetic acid-chelated vacutainers (Becton, Dickinson, and Co.). We isolated, aliquoted, and stored plasma at -80°C after centrifugation at 1200g for ten minutes at 25°C. We collected NP swabs in 1·5mL of RNA Shield Stabilizing Solution (Zymo Research) and stored solution at -80°C. We performed all sample processing under biosafety level 2+ precautions as approved by Stanford University APB-2551.

### SARS-CoV-2 RNA extraction

We extracted RNA from research NP swab and plasma using the QIAamp Viral RNA Mini Kit and QIACube Connect (QIAGEN). We used 140µL of NP swab suspension or plasma as input and eluted RNA in 50µL of elution buffer after lysing for 10 minutes before transferring to QIACube connect.

### SARS-CoV-2 RNA detection and quantification

Multiplexed qPCR and dPCR reactions included the extracted RNA, |Q| Triplex Assay (Combinati), and 4x RT-dPCR MM (Combinati). The |Q| Triplex Assay (Combinati) included primers and probes targeting the N1 and N2 regions of the nucleoprotein gene and the human ribonuclease P gene (RP). We divided the reaction mixture as follows: 10µL for qPCR using the QuantStudio 5 (Applied Biosystems by Thermo Fisher Scientific) and 9µL for dPCR using the array based |Q| (Combinati) (Web Extra Material). Every qPCR and dPCR run included a non-template control (NTC) extracted using the QIACube (NTC-QIA), NTC for PCR, positive extraction control using a combination of the SARS-CoV-2 Specimen positive and negative recombinants (Zeptometrix Corporation), and a positive PCR control with the SARS-CoV-2 Standard from Exact Diagnostics.

We considered a qPCR specimen to be positive if cycle threshold (Ct) for RP, N1, and N2 were all less than 40. For positive samples, we used the lesser (i.e., more readily detected) of N1/N2 Ct for quantitative analysis. We repeated qPCR if the Ct value for RP was greater than 40. We defined a dPCR sample as positive if both N1 and N2 were detected at concentrations of at least 0·23 copies/µL, and used the larger of the two concentrations for quantitative analysis. We set negative qPCR results to 40 (our Ct threshold), and negative dPCR results to zero. We calculated pairwise Pearson’s correlations between measures of qPCR Ct and log-transformed RNA concentrations from dPCR.

Patients with specimens collected on days after enrollment (day zero) typically had specimens collected on day three or seven, not both. Thus, we combined days three and seven (hereafter day 3/7) for purposes of sequential analyses. For patients who had the same test performed on both days, we used the greater of the two values.

### Clinical and laboratory measures

For all participants, we recorded clinical severity of COVID-19 on enrollment and at the time of every additional blood draw, and noted the maximum severity attributable to COVID-19 for 30 days after each patient’s enrollment, using a World Health Organization (WHO) COVID-19 severity scale modified for our institution’s COVID-19 oxygenation protocols: 1 = asymptomatic infection not requiring admission, 2 = symptomatic infection not requiring admission, 3 = admitted without supplemental oxygen, 4 = admitted, requiring oxygen by nasal cannula, 5 = admitted, requiring oxygen by high-flow nasal cannula, 6 = admitted, requiring mechanical ventilation, 7 = admitted, requiring mechanical ventilation *and* vasopressors or renal replacement therapy, 8 = death from COVID-related cause.^20^ We also clustered scores as mild (1-2), moderate (3-4), and severe (5-8). We observed dates associated with initial and maximum WHO severity scores, and each participant’s diagnoses at the time of discharge.

For each participant, we recorded demographic features (age, sex, Hispanic ethnicity), comorbidities (lung disease, cancer, diabetes, immunosuppression, heart disease, hypertension, angiotensin converting enzyme inhibitor [ACE-I] or angiotensin receptor blocker [ARB] use, stroke, dementia, deep venous thrombosis or pulmonary embolus [DVT/PE], chronic kidney disease [CKD], tobacco smoking), initial ED vital signs (systolic [SBP], diastolic [DBP], and mean arterial pressure [MAP], heart rate [HR], respiratory rate [RR], oxygen saturation by pulse oximetry [SpO2], temperature), presence of pneumonia on chest X-ray or computed tomography, patient-reported symptoms (fever, chills, cough, sore throat, congestion, shortness of breath, chest pain, myalgias, nausea, vomiting, diarrhea, loss of taste, loss of smell, confusion, headache), and laboratory values (leukocyte count, absolute lymphocyte count, haemoglobin, platelet count, D-dimer, fibrinogen, prothrombin time [PT], partial thromboplastin time [PTT], erythrocyte sedimentation rate [ESR], C-related peptide [CRP], procalcitonin, lactate dehydrogenase [LDH], ferritin, troponin, lactate, sodium, potassium, chloride, bicarbonate, blood urea nitrogen [BUN], creatinine, calcium, magnesium, glucose, bilirubin, aspartate aminotransferase [AST], alanine aminotransferase [ALT], alkaline phosphatase).

We dichotomised vital signs as follows: MAP was low if below 88mmHg (the 25^th^ percentile for enrolled patients), high if above 140mmHg (75^th^ percentile). We did not use a typical clinical definition of hypotension such as MAP below 65mmHg because no patients met this threshold at the time of study enrollment. RR was low if less than 8, high if greater than 20. SpO2 was low if less than 95% (the 25^th^ percentile). Temperature was high if 38°C or greater, low if less than 36°C. We dichotomised laboratory values (high or low) based on our laboratory’s normal ranges. For the purpose of predictive models, missing dichotomised laboratory values were imputed nonparametrically for each participant with a Random Forests model, using 100 trees, and 10 iterations (R package *missForest* version 1·4).

### Defining extrapulmonary complications

We created binary indicators for whether each participant had any of six EPCs during their encounter. Patients were considered to have *neurologic* involvement if they were diagnosed with one or more of: acute stroke, encephalitis, meningitis, neuroinflammatory disease, delirium. *Cardiovascular* involvement included myocardial injury, acute coronary syndrome, cardiomyopathy, acute cor pulmonale, arrhythmia, or cardiogenic shock. *Renal* involvement was defined by acute kidney injury. Transaminitis or hyperbilirubinemia constituted *hepatobiliary* involvement. *Hematologic* involvement included deep vein thrombosis, pulmonary embolus, myocardial infarction, acute stroke, acute limb ischemia, mesenteric ischemia, or catheter-related thrombosis. Patients were considered to have *immunologic* involvement if they received a diagnosis of sepsis, septic shock, multi-organ failure, or secondary bacterial infection.

### Characterising associations between RNAaemia and clinical severity

We calculated mean maximum WHO severity scores for patients who were initially RNAaemic and non-RNAaemic, and compared mean scores with a two-sample t-test. Among RNAaemic patients, we calculated Pearson’s correlation between log-transformed concentration of RNA in plasma, and maximal clinical severity (WHO 1-8). We calculated proportions of RNAaemic and non-RNAaemic patients who manifested mild (WHO 1-2), moderate (WHO 3-4), and severe (WHO 5-8) disease, who were admitted to the hospital, who manifested EPCs, and who worsened after presentation (i.e., had a maximum WHO score exceeding WHO score at enrollment). We compared the proportions of RNAaemic and non-RNAaemic patients in each of those categories using chi-squared tests with continuity corrections. We calculated the odds ratio for clinical deterioration, by RNAaemia on enrollment (using the odds ratio for consistency with logistic regression results), and calculated exact 95% confidence intervals.^21^ We compared median length-of-hospitalisation in days, and median degree of clinical worsening (difference between initial and maximum WHO score), for RNAaemic and non-RNAaemic patients, using the Wilcoxon rank-sum test with continuity correction to compare these distributions.

### Predictive models for severe disease, extrapulmonary complications, and RNAaemia

We developed a predictive model for severe (WHO 5-8) disease, based on data available upon patient presentation. We included the following variables as potential predictors of severe disease: demographic features, comorbidities, binary indicators of abnormal ED vital signs, pneumonia on chest X-ray or CT, patient-reported symptoms, and binary indicators of abnormal lab values (described above). Because therapies such as Remdesivir and dexamethasone were generally initiated after ED presentation, and on the basis of oxygen requirements (the main constituent of the WHO severity score), we did not include specific treatments in our predictive models.

To prevent overfitting, given a large number of potential predictors compared to the number of patients, we selected variables via elastic net regularisation (*glmnet* 4·0 in *R*), using logistic models and 10-fold cross-validation, selecting the regularisation parameter λ minimising mean cross-validated error. We then used the selected variables in a logistic model, and estimated odds ratios and 95% confidence intervals for prediction of severe disease, for each of the selected features. We calculated mean cross-validated area under the receiver-operating characteristic curve (AUROC) of the resulting model.

We predicted the presence of EPCs in analogous fashion, excluding the patient-reported symptoms potentially constitutive of EPCs as defined above, and excluding laboratory markers, many of which were used in definition of EPCs. We again selected variables via cross-validated elastic-net regularisation, estimated odds ratios for the most robust predictors of EPCs, and characterised overall predictive accuracy by AUROC.

Finally, we predicted the presence of RNAaemia in analogous fashion, using as potential predictors demographic features, comorbidities, and symptoms, but excluding radiographic and laboratory findings.

## Results

### Patient characteristics on enrollment

We enrolled 191 COVID-19 positive ED patients, all of whom had plasma sampled on day of enrollment (day zero). Some patients had additional NP or plasma samples collected at one or more of days: three, seven, 30. 49·2% (94/191) of participants were women. Median age was 47 years (IQR 34 - 61). Patients had a median of one comorbidity (IQR 0-3) from the following list: lung disease, cancer, diabetes, immunosuppression, heart disease, hypertension, ACE-I/ARB use, stroke, dementia, DVT/PE, CKD. Patients reported a median of four (IQR 2-6) symptoms from the following list: fever, chills, cough, sore throat, congestion, shortness of breath, chest pain, myalgia, nausea/vomiting/diarrhea (any), loss of taste, loss of smell, confusion, headache. Patient characteristics at enrollment are summarised in Table 1.

**Table 1.** Patient characteristics on enrollment.

| Characteristic | Value |
| --- | --- |
| N | 191 |
| Female | 49.2% (94/191) |
| Median age (IQR) | 47 (IQR 34 - 61) |
| **Medical history:** |  |
| Lung disease | 12.6% (24/191) |
| Cancer | 13.6% (26/191) |
| Diabetes | 26.7% (51/191) |
| Immunosuppression | 7.3% (14/191) |
| Heart disease | 11.0% (21/191) |
| Hypertension | 36.6% (70/191) |
| ACE/ARB use | 18.3% (35/191) |
| Stroke | 4.2% (8/191) |
| Dementia | 4.7% (9/191) |
| DVT/PE | 5.8% (11/191) |
| Chronic kidney disease | 9.9% (19/191) |
| Smoking | 20.9% (40/191) |
| **Symptoms on presentation:** |  |
| Fever | 64.4% (123/191) |
| Chills | 31.4% (60/191) |
| Cough | 67.5% (129/191) |
| Sore throat | 16.2% (31/191) |
| Congestion | 8.4% (16/191) |
| Shortness of breath | 63.4% (121/191) |
| Chest pain | 34.6% (66/191) |
| Myalgia | 34.6% (66/191) |
| Nausea/vomiting/diarrhea | 40.8% (78/191) |
| Loss of taste | 38.7% (74/191) |
| Loss of smell | 27.2% (52/191) |
| Confusion | 2.6% (5/191) |
| Headache | 26.2% (50/191) |

ACE = Angiotensin-converting enzyme inhibitor, ARB = Angiotensin receptor blocker, DVT = deep vein thrombosis, PE = pulmonary embolus.

### SARS-CoV-2 RNA prevalence by sample type, method, and day of collection

dPCR was more sensitive than qPCR for the detection of RNAaemia, detecting RNAaemia in 23·0% (44/191) of patients on day zero, compared to 1·4% (2/147) for qPCR. On day three, dPCR detected RNAaemia in 13·3% (6/45) of specimens, compared to zero for qPCR. On day seven, dPCR detected RNAaemia in 6·8% (3/44), compared to zero for qPCR. At day 30, neither dPCR nor qPCR detected RNAaemia in the 32 specimens tested. We describe the analytical performance of the dPCR assay in the Web Extra Material.

We observed a modest negative correlation (*r* = -0·30) between qPCR Ct values and dPCR RNA concentrations for the same plasma specimens (Figure 1). Notably, the correlation between NP and plasma dPCR values across 48 paired specimens was very weak *(r* = 0·16). Plasma RNA by dPCR on day zero was moderately correlated with the same measure on day 3/7 (*r =* 0·42).

**Figure 1.**
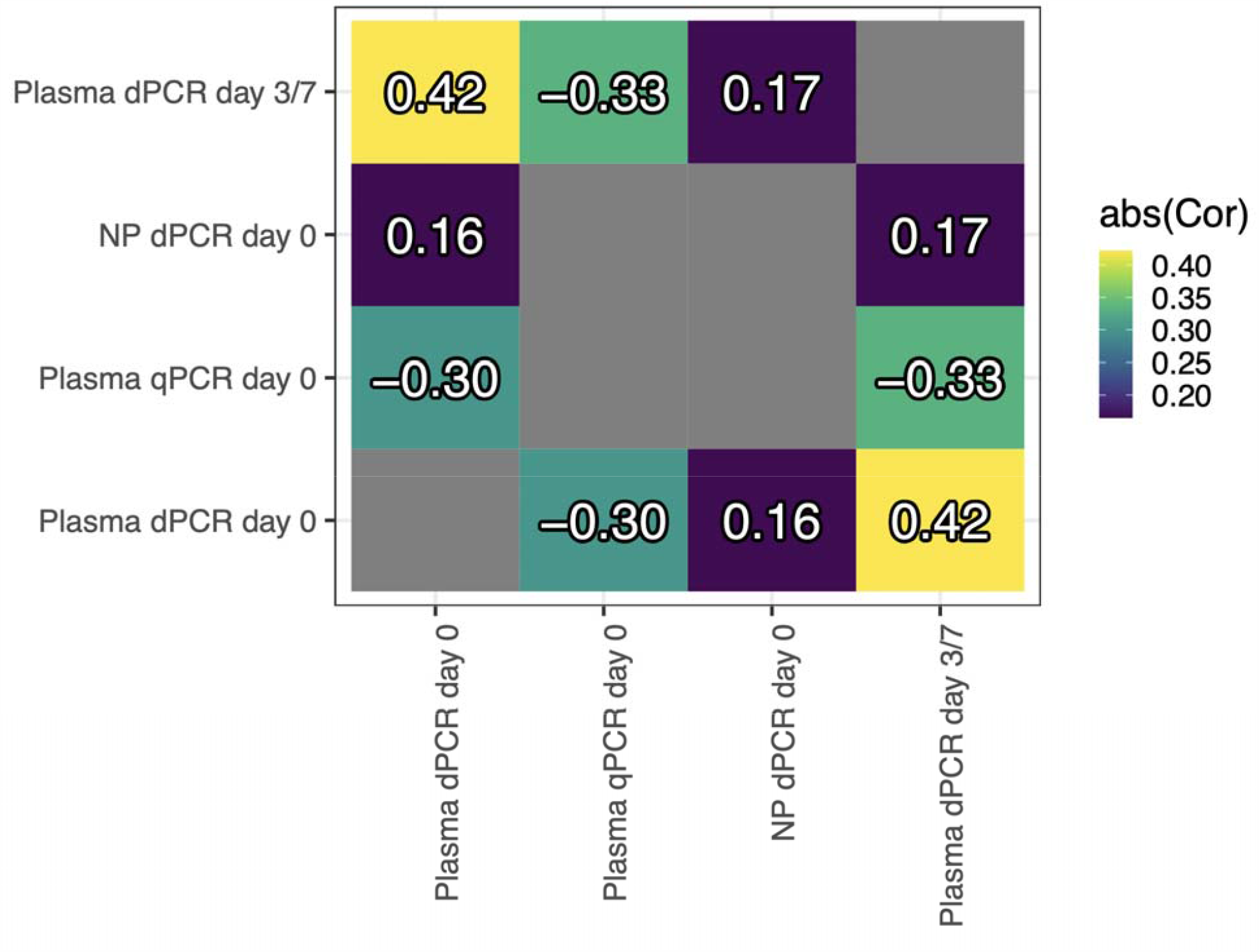
Pairwise Pearson’s correlations between measures of nasopharyngeal (NP) and plasma SARS-CoV-2 RNA load. Colors reflect absolute pairwise correlation, as qPCR cycle thresholds are expected to be inversel proportional to SARS-CoV-2 RNA concentrations as measured by dPCR. Plasma RNA concentration by dPCR at enrollment (“Plasma dPCR day 0”) is modestly negatively correlated (*r* = -0·30) with qPCR Ct on the same specimen, moderately correlated with plasma dPCR on day 3/7 (*r* = 0·42), and poorly correlated with RNA concentration in the nasopharynx (*r* = 0·16), suggesting that RNAaemia is weakly related to nasopharyngeal viral load. NP = nasopharyngeal swab. qPCR = quantitative PCR. dPCR = digital PCR.

### Persistence of RNAaemia

We classified the maximum clinical severity for each enrolled patient as mild (54), moderate (104), or severe (33) (Figure 2). Of the 44 patients RNAaemic by dPCR on day zero, 27 had additional draws on days three, seven, or 30. Of these, 92·6% (25/27) had highest RNA on day zero, 22·2% (6/27) were persistently RNAaemic at day 3/7, and 7·4% (2/27) had rising RNA by day 3/7 after enrollment. 50 patients with positive screening swabs who were not RNAaemic on day zero had plasma drawn again on day 3/7. Of these, only 2·0% (1/50) became RNAaemic on day 3/7.

**Figure 2.**
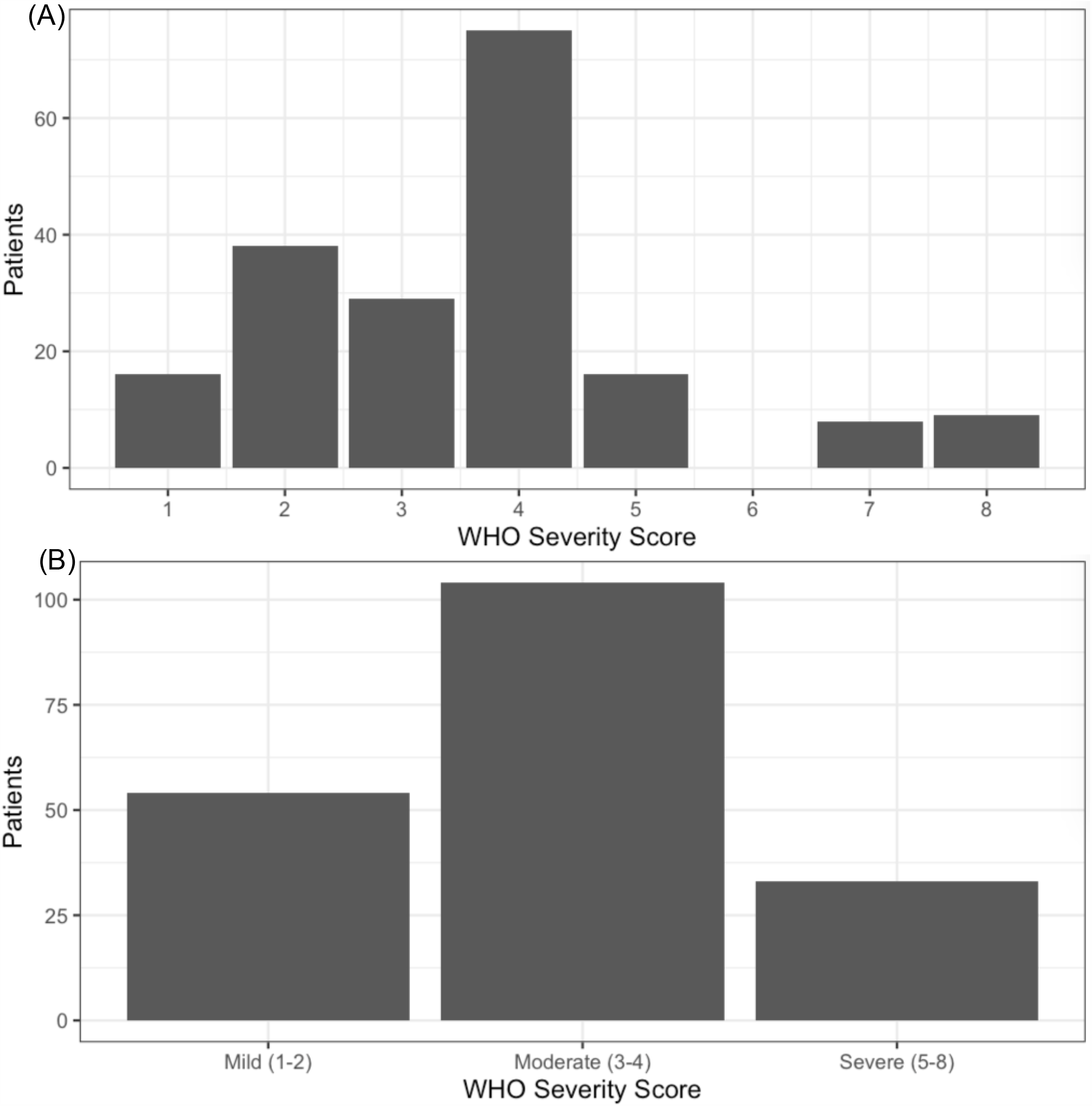
Distribution of discrete and binned WHO severity scores. We classified the maximum severity of 14 SARS-CoV-2 presentations using a modified WHO score, as follows: 1 = asymptomatic infection, 2 = symptomatic infection not requiring admission, 3 = admitted without supplemental oxygen, 4 = admitted, requiring oxygen by nasal cannula, 5 = admitted, requiring oxygen by high-flow nasal cannula, 6 = admitted, requiring mechanical ventilation, 7 = admitted, requiring mechanical ventilation and vasopressors or renal replacement therapy, 8 = deat from COVID-related cause. **A**. Distribution of WHO scores. **B**. Distribution of binned (mild, moderate, severe) scores.

### RNAaemia and severity of disease

RNAaemic patients had higher mean clinical severity (4·80) than non-RNAaemic patients (3·24, difference = 1·56 [95% CI, 1·00 - 2·11]). 40·9% of RNAaemic patients developed severe disease, compared to 10·2% of non-RNAaemic patients (difference = 30·7% [95% CI of difference, 13·9% - 47·5%]). Conversely, 4·5% of initially RNAaemic patients had mild disease, compared to 35·4% of non-RNAaemic patients (difference = 30·8% [95% CI of difference, 19·5% - 42·2%]) (Figure 3A). Among patients with detectable RNAaemia at time of enrollment (n=44), patients with higher plasma RNA concentrations manifested more severe disease (*r* = 0·47 [95% CI, 0·20 - 0·67]) (Figure 3B). Severity trended higher in persistently RNAaemic patients, compared to patients RNAaemic at day zero but not at day 3/7 (mean WHO score 6·5 vs 5·0), but the difference was not significant (95% CI of difference, -0·58 - 3·58).

**Figure 3.**
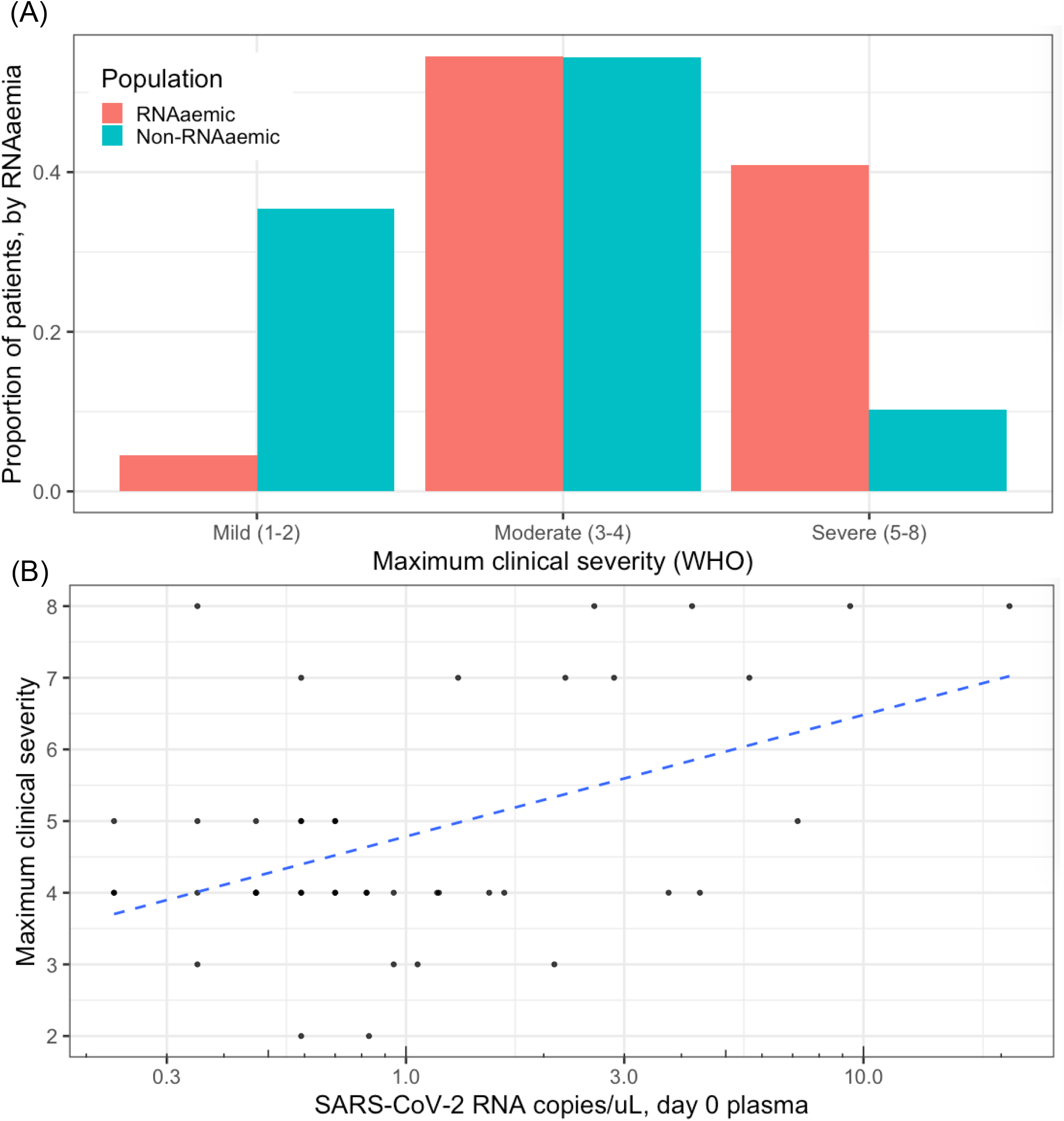
SARS-CoV-2 RNAaemia and clinical severity. **A**. RNAaemic patients had higher mean maximum WHO scores (4·80) than non-RNAaemic patients (3·24, difference = 1·56 [95% CI of difference, 1·00 - 2·11]). 40·9% of RNAaemic patients developed severe disease, compared to 10·2% of non-RNAaemic patients (difference = 30·7% [95% CI of difference, 13·9% - 47·5%]). 4·5% of initially RNAaemic patients had mild disease, compare to 35·4% of non-RNAaemic patients (difference = 30·8% [95% CI of difference, 19·5% - 42·2%]). The same proportion (54·5%) of both RNAaemic and non-RNAaemic patients had disease of moderate severity. **B**. Among patients with detectable RNAaemia at time of enrollment (n=44), patients with higher plasma RNA concentrations manifested more severe disease (*r* = 0·47 [95% CI, 0·20 - 0·67]). RNA concentrations in RNAaemic patients were distributed approximately log-normally, so were log-scaled for depiction and calculation of correlation. Dashed blue line shows linear correlation between log-scaled plasma RNA concentration and maximum clinical severity.

90·9% (40/44) of RNAaemic patients, and 70·1% (103/147) of non-RNAaemic patients required hospital admission (difference = 20·8% [95% CI, 8·1% - 33·6%]). Among admitted patients, RNAaemic patients had longer median length-of-stay (7·6 vs 5·1 days, *p* < 0·01, Wilcoxon rank-sum test with continuity correction).

In an elastic-net regularised, cross-validated logistic model of severe (WHO 5-8) disease, the significant predictors of severe disease were tobacco smoking, low SpO2 on ED arrival, and RNAaemia (OR of RNAaemia for severe disease = 6·72 [95% CI, 2·45 – 19·79]). The overall predictive performance of the model was good, with mean cross-validated AUROC of 0·82 (Table 2).

**Table 2.** Prediction of severe disease. Potential predictors of severe (WHO 5-8) disease included: demographic features (age 60+ or 80+, sex), past medical history features (lung disease, cancer, diabetes, immunosuppression, heart disease, hypertension, angiotensin converting enzyme inhibitor or angiotensin receptor blocker use, stroke, dementia, deep venous thrombosis or pulmonary embolus, chronic kidney disease, tobacco smoking, obesity), binary indicators of abnormal ED vital signs (low or high mean arterial pressure, low or high heart rate, low or high respiratory rate, low oxygen saturation, low or high temperature), pneumonia on chest X-ray or CT, patient-reported symptoms (fever, chills, cough, sore throat, congestion, shortness of breath, chest pain, myalgias, nausea/vomiting/diarrhea, loss of taste, loss of smell, confusion, headache), and binary indicators of abnormal lab values (high or low leukocyte count, low absolute lymphocyte count, low haemoglobin, low or high platelet count, high D-dimer level, high fibrinogen level, low fibrinogen level, high prothrombin time, high partial thromboplastin time, high C-related peptide level, high lactate dehydrogenase level, high ferritin level, high troponin level, high lactate level, high or low sodium level, high or low potassium level, high or low chloride level, high or low bicarbonate level, high blood urea nitrogen level, high creatinine level, high or low calcium level, high or low magnesium level, high or low glucose level, high bilirubin level, high aspartate aminotransferase level, high alanine aminotransferase level, high alkaline phosphatase level). To prevent over-fitting, predictors were selected via elastic net regression of severe disease on these features with 10-fold cross-validation, selecting the regularisation parameter λ minimizing mean cross-validated error, and yielding the features in the table above. In a logistic model regressing severe disease on these features, significant predictors of severe disease included: tobacco smoking, low oxygen saturation (SpO2), and RNAaemia. RNAaemia was associated with 6·7 times the odds of severe disease, adjusting for other features selected by elastic net penalised regression, an association comparable in magnitude to the association of hypoxia on initial presentation with eventual severe disease. Mean cross-validated area under the receiver-operating characteristic curve (AUROC) of the model in predicting severe disease was 0·82.

|  | OR (95% CI) |
| --- | --- |
| PMH: DM | 1.51 (0.51 - 4.45) |
| **Smoker** | **3.13 (1.08 - 9.38)** |
| ED: MAP low | 2.59 (0.92 - 7.47) |
| **ED: SpO2 low** | **5.36 (2.03 - 15.07)** |
| ALC low | 3.12 (1.00 - 9.8) |
| Lactate high | 3.90 (0.63 - 22.91) |
| Glucose high | 2.58 (0.92 - 7.30) |
| **RNAemic** | **6.72 (2.45 - 19.79)** |
| N | 191 |
| AIC | 134.74 |
| AUROC | 0.82 |

### Dynamics of infection through course of illness

27 patients had RNAaemia at enrollment and one or more subsequent plasma samples. A majority (14/27) of these patients had undetectable RNAaemia by day ten from symptom onset, while the same proportion took 16 days to reach maximum severity, and 33 days until resolution of symptoms (Figure 4). Of these 27 patients, 2 had mild disease at enrollment, 20 had moderate disease, and 5 had severe disease. Through the disease course, 17 improved to mild severity, six remained severe, and three died in the hospital.

**Figure 4.**
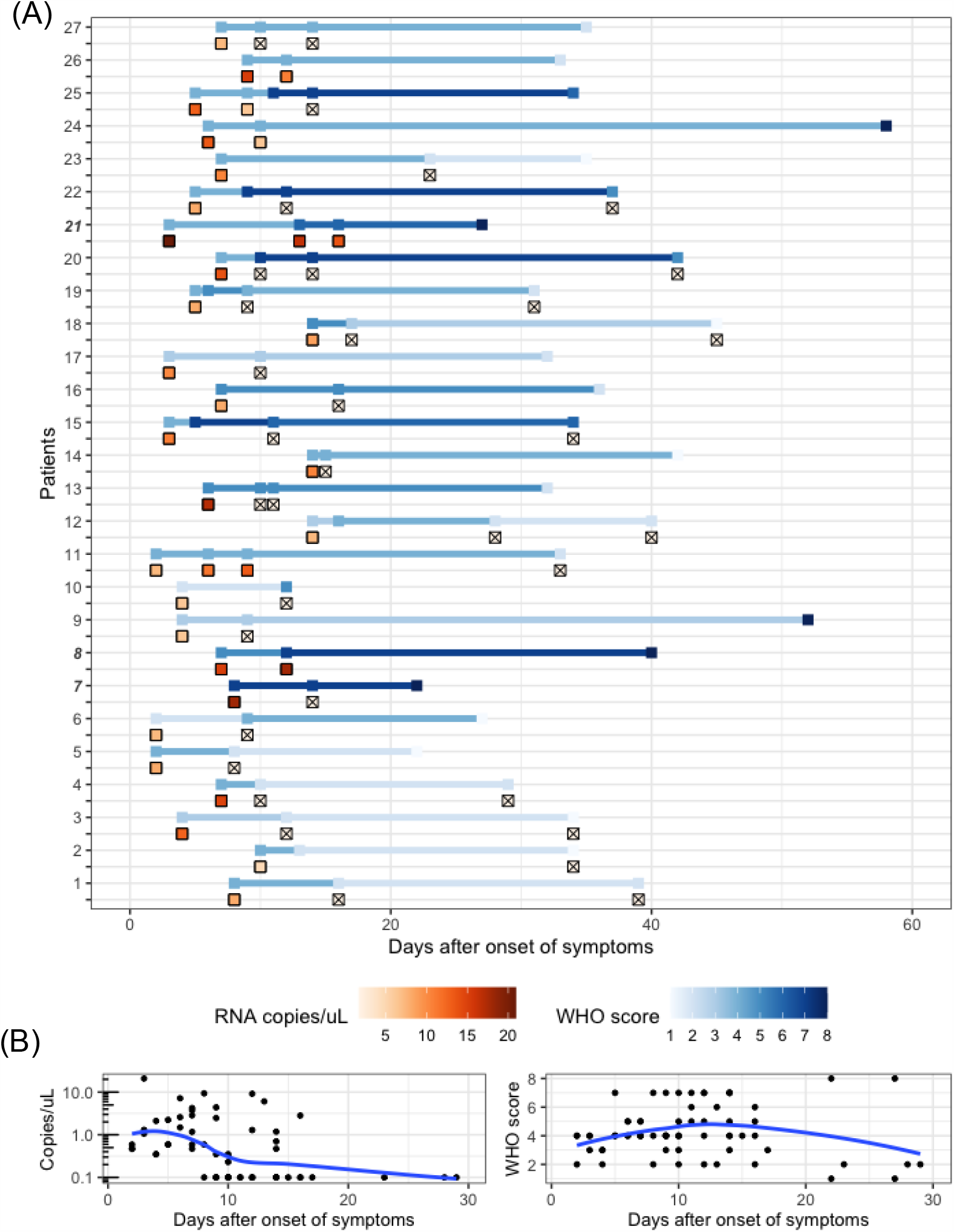
Dynamics of SARS-CoV-2 RNAaemia and clinical severity, by modified WHO score. **A**. Serial plasma SARS-CoV-2 RNA concentrations and WHO scores for each of the 27 patients with longitudinal samples. Plasma RNA concentration (red gradient) and WHO scores (blue gradient) are shown with respect to the number of days since the reported onset of symptoms (not date of study enrollment) for each patient. Patients who died in the hospital are highlighted in bold and italics. Specimens with undetectable RNAaemia are represented as Ill. Most (14/27) patients had undetectable RNAaemia by day 10, while the same proportion took 16 days to reach maximum severity, and 33 days for resolution of symptoms. **B**. Aggregate RNA and clinical dynamics in the 30 days following onset of symptoms. Loess regression curves represent trends in RNA and clinical dynamics. RNAaemia peaked 3 days after symptom onset, while clinical severity peaked at 14 days.

In the total study population (i.e., not restricted to those with serial plasma samples), 77·0% (147/191) of patients manifested their maximum clinical severity on the day of study enrollment (i.e., when patients presented to the ED), while 23·0% (44/191) worsened 24 hours or more after enrollment (Figure 5). 36·4% (16/44) of initially RNAaemic patients, and 19·0% (28/147) of non-RNAaemic patients worsened in severity after initial presentation (difference = 17·4% [95% CI of difference, 0·3% - 34·4%], OR 2·43 [95% CI, 1·07 - 5·38]). RNAaemic patients worsened by a median of three points on the modified WHO scale, compared to one point for non-RNAaemic patients (*p =* 0·02, Wilcoxon rank-sum test with continuity correction).

**Figure 5.**
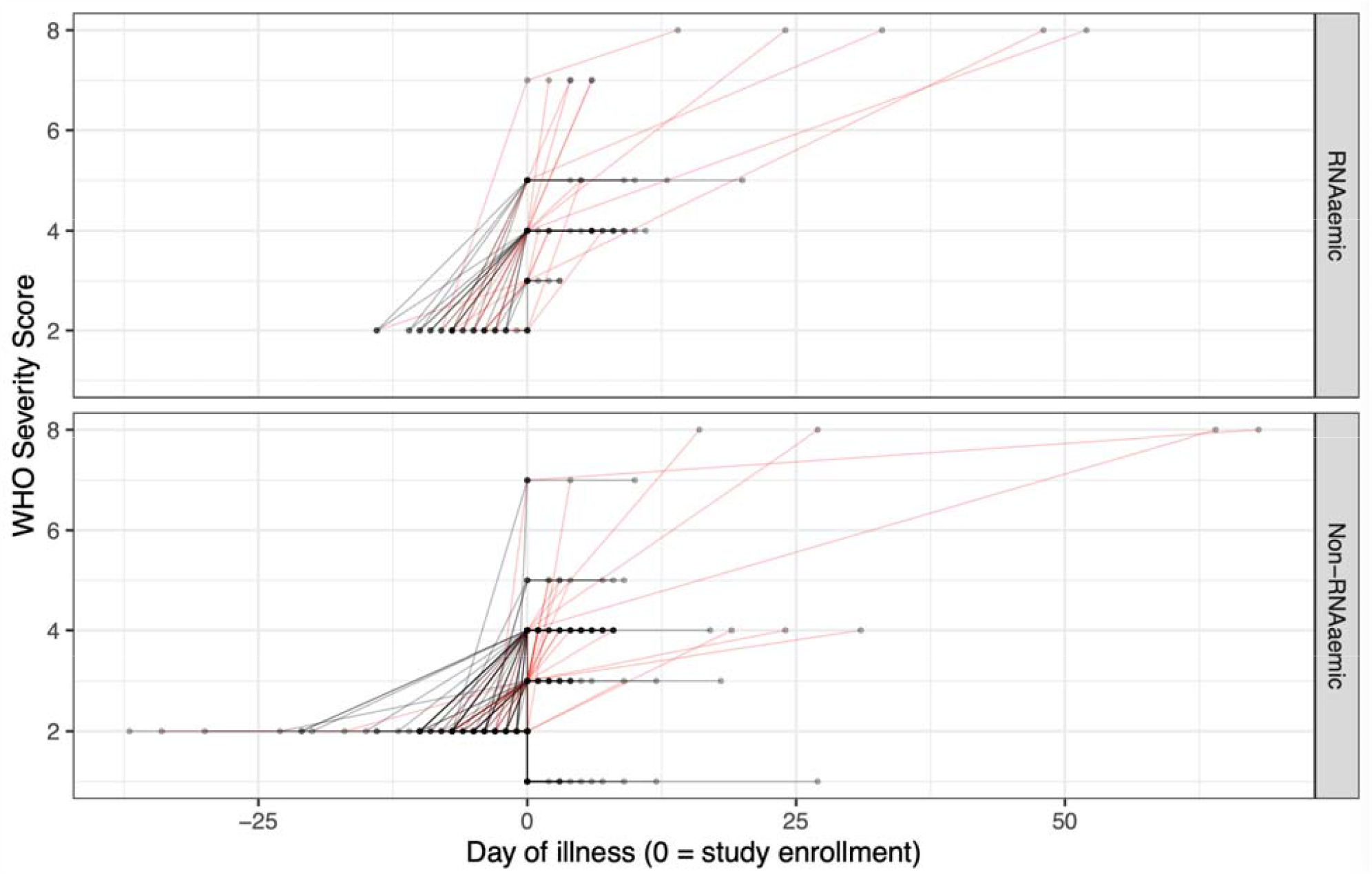
Trajectories of patient severity, by RNAaemia on initial presentation. 36·4% (16/44) of initially RNAaemic patients, and 19·0% (28/147) of non-RNAaemic patients worsened in severity after initial presentatio (difference = 17·4% [95% CI of difference, 0·3% - 34·4%]). RNAaemic patients worsened by a median of three points on the modified WHO scale, compared to one point for non-RNAaemic patients (*p =* 0·02, Wilcoxon rank-sum test with continuity correction). Day zero represents day of patient enrollment. Values prior to day zero are based on patient’s first reported day of symptoms. Values after day zero are based on the date of each patient’s maximum WHO score. Red trajectories are those that increase in severity after presentation, by modified WHO score.

### RNAaemia predicts extrapulmonary complications

56·8% (25/44) of RNAaemic patients developed one or more extrapulmonary complications, compared to 30·6% (45/147) those non-RNAaemic (difference in proportions = 26·2% [95% CI, 8·3% - 44·1%]) (Figure 6). RNAaemic patients tended toward higher rates of EPCs across systems, though only differences in rates of hepatobiliary, haematologic, and immunologic complications were individually statistically significant at *p* < 0·05 (chi-squared tests for equality of proportions with continuity correction).

**Figure 6.**
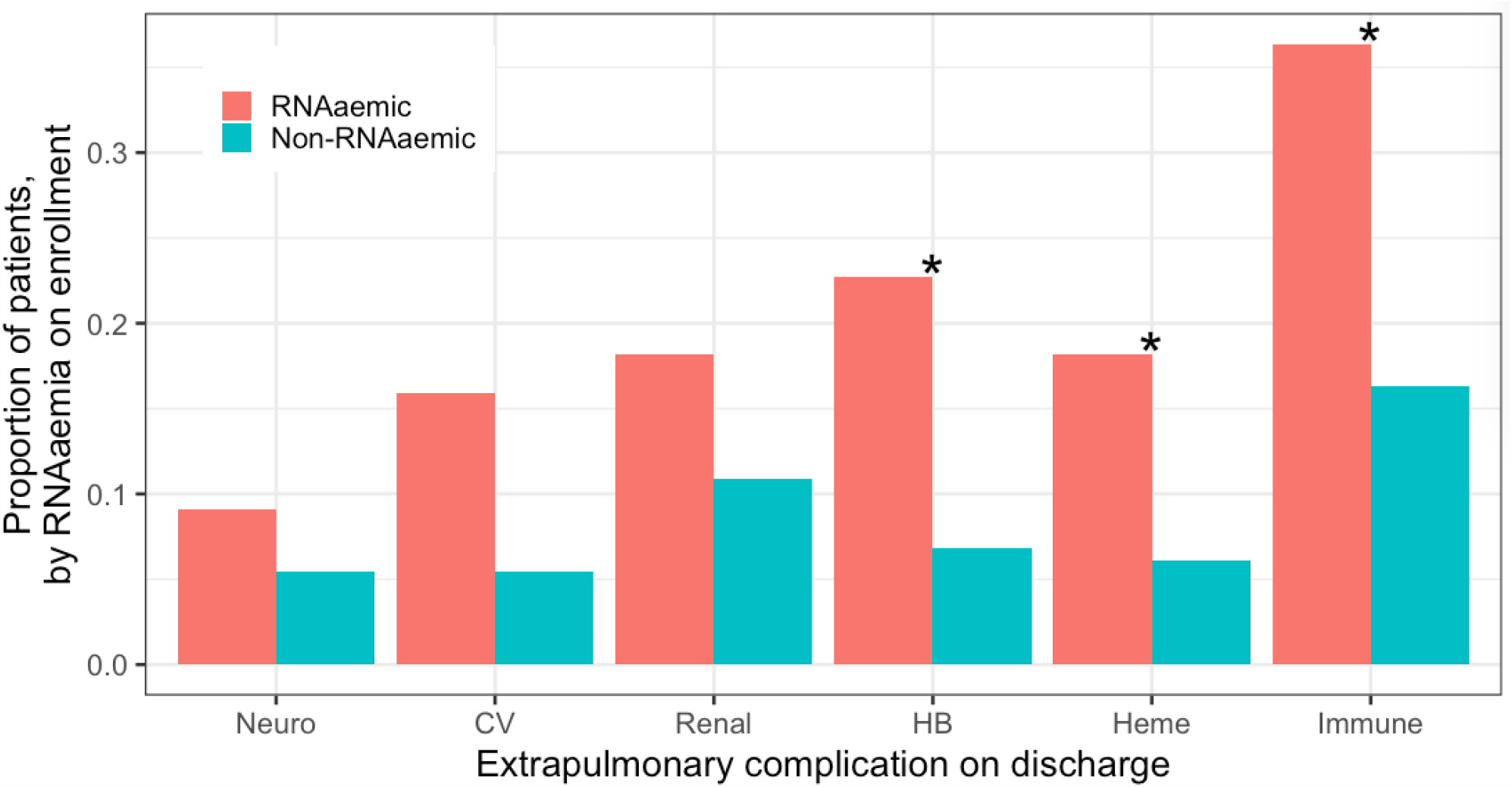
Presence of extrapulmonary complications, by RNAaemia. 56·8% (25/44) of patients RNAaemic on enrollment patients developed one or more extrapulmonary complications by hospital discharge, compared to 30·6% (45/147) of non-RNAaemic patients (difference in proportions = 26·2% [95% CI, 8·3% - 44·1%]). RNAaemi patients tended toward higher rates of extrapulmonary complications across systems, though only differences i rates of hepatobiliary (HB), haematologic, and immunologic complications were individually statistically significant at *p* < 0·05 (chi-squared tests for equality of proportions with continuity correction). *CV* = cardiovascular, *HB =* hepatobiliary.

In an elastic-net regularised, cross-validated logistic model, significant predictors that a patient would manifest one or more EPCs were: chronic kidney disease, obesity, and RNAaemia (OR 2·81 [95% CI, 1·26 – 6·36]). The overall predictive performance of the model was fair, with mean cross-validated AUROC of 0·73 (Table 3).

**Table 3.** Prediction of extrapulmonary complications. Potential predictors of extrapulmonary complications (EPCs) included: demographic features (age 60+ or 80+, sex), past medical history features (lung disease, cancer, diabetes, immunosuppression, heart disease, hypertension, angiotensin converting enzyme inhibitor or angiotensin receptor blocker use, stroke, dementia, deep venous thrombosis or pulmonary embolus, chronic kidney disease, tobacco smoking, obesity), binary indicators of abnormal ED vital signs (low or high mean arterial pressure, low or high heart rate, low or high respiratory rate, low oxygen saturation, low or high temperature), pneumonia on initial chest X-ray or CT, and patient-reported symptoms on enrollment excluding those constitutive of extrapulmonary diagnosis (fever, chills, cough, sore throat, congestion, shortness of breath, chest pain, myalgias). Laboratory values were not included as many were constitutive of extrapulmonary diagnoses. To prevent over-fitting, predictors were selected via elastic net regression of EPC (1 if a patient had one or more EPC, 0 if none) on these features with 10-fold cross-validation, selecting the regularization parameter λ minimising mean cross-validated error, and yielding the features in the table above. In a logistic model regressing EPC on these features, significant predictors of EPC included: chronic kidney disease, obesity (BMI>30), and RNAaemia. RNAaemia was associated with 2·8 times the odds of EPC, comparable in magnitude to the association between obesity and development of EPC. Mean cross-validated area under the receiver-operating characteristic curve (AUROC) of the model in predicting EPC was 0·73.

|  | OR (95% CI) |
| --- | --- |
| Age: 80+ | 2.27 (0.49 - 9.92) |
| PMH: Heart | 2.13 (0.63 - 7.41) |
| PMH: HTN | 1.74 (0.81 - 3.69) |
| PMH: Dementia | 3.60 (0.58 - 25.33) |
| **PMH: CKD** | **4.56 (1.36 - 17.27)** |
| Smoker | 1.88 (0.80 - 4.42) |
| **Obese** | **2.64 (1.23 - 5.84)** |
| ED: RR high | 1.63 (0.79 - 3.35) |
| ED: SpO2 low | 1.34 (0.60 - 2.93) |
| **RNAemic** | **2.81 (1.26 - 6.36)** |
| N | 191 |
| AIC | 222.25 |
| AUROC | 0.73 |

### Prediction of RNAaemia on presentation

We sought to determine whether the patients with RNAaemia on presentation could be predicted on the basis of their demographics, comorbidities, symptoms, and ED vital signs. In an analogous regularised, cross-validated logistic model (Table S1), significant predictors of RNAaemia on presentation were limited to cough and hypoxia, though the overall predictive performance of the model was poor (mean cross-validated AUROC 0·66).

## Discussion

The pathogenesis of COVID-19, its temporal dynamics, and the determinants of disease severity and extrapulmonary complications are incompletely understood. We explored the performance and clinical utility of dPCR in quantifying SARS-CoV-2 RNA in the nasopharynx and plasma, and characterised the relationships between RNAaemia and disease severity, clinical deterioration, and extrapulmonary complications. Array-based dPCR was much more sensitive than qPCR for the detection of SARS-CoV-2 in plasma, where mean concentration of viral RNA was three orders of magnitude less than in the nasopharynx. RNAaemia manifests early in the course of illness, while clinical manifestations peak later and are more prolonged. RNAaemia at presentation predicts severe disease, ongoing clinical deterioration, and specific extrapulmonary complications.

We found dPCR to be markedly more sensitive than qPCR even while using more stringent detection criteria (both N1 and N2 >=0·23 copies/µL) than other studies (e.g., either N1 or N2 >=0·1 copies/µL).^8^ dPCR was also more consistent in multiplex detection of both regions (N1 and N2) of the SARS-CoV-2 nucleocapsid gene, likely related to the partition format in dPCR reducing preferential amplifications often observed in bulk PCR.^22^ Moreover, a dPCR platform based on microwell array enhances partition consistency, minimises dead volume for improved sensitivity, and has a workflow identical to qPCR, making it suitable for incorporation into existing clinical workflows. Early in the course of an outbreak, before viral RNA standard curves are widely available, dPCR is a natural choice for detecting a novel pathogen.

RNAaemia on presentation was a major predictor of both severe disease and extrapulmonary complications, after accounting for demographics, comorbidities, symptoms, vital signs, and a host of laboratory markers. Moreover, RNAaemic patients were more likely than non-RNAaemic patients to worsen after presentation, and RNAaemic patients who worsened did so by a greater degree. Previous studies have associated RNAaemia with disease severity and mortality.^8,10,23^ Reported associations between RNAaemia and extrapulmonary involvement are more varied.^2,24,25^ In addition to characterising the temporal dynamics of clinical deterioration, we included a more comprehensive scope of potential confounders than previous studies.^8^ We also use cross-validation not only for model selection, but to assess the relative predictability of clinical severity (good, AUROC 0·82), extrapulmonary complications (fair, AUROC 0·73), and RNAaemia itself (poor, AUROC 0·66). The poor predictability of RNAaemia from patient features observable on initial presentation, and the weak correlations between NP and plasma measurements of SARS-CoV-2 RNA, suggest that RNAaemia is not simply a consequence of sufficient viral load at the typical site of inoculation, but may instead signal unique pathophysiologic and prognostic features.^6,26,27^

SARS-CoV-2 RNAaemia might arise from spillage from the respiratory tract, or from active viral replication in vascular endothelial or perivascular cells.^28,29^ Whether RNAaemia represents intact, replicating virus cannot be directly determined from our data, and an attempt to culture SARS-CoV-2 from serum with low RNA levels was not successful.^30^ SARS-CoV-1, however, has been found to replicate in circulating lymphocytes, monocytes, macrophages, and dendritic cells.^31–33^ The RNAaemia kinetics we observed follow a typical viral kinetic pattern, with high peak viral load early in the infection, followed by rapid decay (likely reflecting the innate immune response), before a slower clearance (from acquired immunity).^34^ RNAaemia prior to symptom onset has been anecdotally reported; more data is needed to better assess potential pre-symptomatic dynamics.^30^ Since our findings suggest that RNAaemia on presentation reflects the likelihood of subsequent disease progression, early testing for RNAaemia could prove useful for the targeted initiation and monitoring of antiviral therapies.^34^

The association we observed between RNAaemia and EPCs (which we defined conservatively based on diagnoses at discharge, rather than surrogate biomarkers alone), is stronger than in some previous reports.^25^ Extrapulmonary injury could result from direct viral toxicity, endothelial cell damage and thromboinflammation, dysregulation of the immune response, or dysregulation of the renin–angiotensin–aldosterone system.^2^ Transaminitis, a common hepatobiliary complication we observed in RNAaemic patients, might result from direct hepatocellular injury by SARS-CoV-2, from cytokine storm and hypoxia-associated metabolic derangement, or from drug-induced liver injury (particularly secondary to investigational agents such as remdesivir, lopinavir, and tocilizumab). The trend we observed toward higher incidence of acute kidney injury in RNAaemic patients is consistent with prior evidence for renal tropism.^12^

We found that SARS-CoV-2 RNAaemia at the time of initial ED presentation is a robust predictor of patients’ eventual clinical severity and EPCs. Despite the limited generalisability of our single centre study, the substantial predictive value of RNAaemia in multiple aspects of the disease course suggests a role for plasma dPCR in triage and disposition. Since we use a measure of severity based primarily on oxygen requirements (i.e., the modified WHO score), and since many COVID-19 therapies are initiated on the basis of oxygen requirements, RNAaemia on presentation might serve to direct early initiation of appropriate therapies for the patients most likely to deteriorate.

## Supporting information

Web Extra Material

## Data Availability

De-identified study data are presented as online datasets.

## Contributors

SY conceived the study. SY, AJR, CAB, KN, ALB, RO, EA, JAN, JVQ established the Stanford COVID-19 Biobank. EJZ enrolled patients and compiled clinical data. NR, MMH, KT, HN, LJ, PH collected and processed the PCR data. DK did the modelling and statistical analysis. NR and DK did the longitudinal analysis. DK and NR drafted the manuscript and produced the figures. All authors contributed to writing the manuscript and approved the final version.

## Data sharing

De-identified study data are presented as online datasets.

## Acknowledgements

Additional author members of the Stanford COVID-19 Biobank Study Group include: Rosen Mann, Anita Visweswaran, Thanmayi Ranganath, Jonasel Roque, Monali Manohar, Hena Naz Din, Komal Kumar, Kathryn Jee, Brigit Noon, Jill Anderson, Bethany Fay, Donald Schreiber, Nancy Zhao, Rosemary Vergara, Julia McKechnie, Aaron Wilk, Lauren de la Parte, Kathleen Whittle Dantzler, Maureen Ty, Nimish Kathale, Arjun Rustagi, Giovanny Martinez-Colon, Geoff Ivison, Ruoxi Pi, Maddie Lee, Rachel Brewer, Taylor Hollis, Andrea Baird, Michele Ugur, Drina Bogusch, Georgie Nahass, Kazim Haider, Kim Quyen Thi Tran, Laura Simpson, Michal Tal, Iris Chang, Evan Do, Andrea Fernandes, Allie Lee, Neera Ahuja, Theo Snow, James Krempski.

