## Supplementary material for "SARS-CoV-2 RNAaemia predicts clinical deterioration and extrapulmonary complications from COVID-19": Web Extra Material

**Web Extra Material: SARS-CoV-2 RNAaemia predicts clinical deterioration and extrapulmonary complications from COVID-19.**

Nikhil Ram-Mohan*, David Kim*, Elizabeth J Zudock, Marjan M Hashemi, Kristel Tjandra, Angela J Rogers, Catherine A Blish, Kari C. Nadeau, Jennifer A Newberry, James V Quinn, Ruth O’Hara, Euan Ashley, Hien Nguyen, Lingxia Jiang, Paul Hung, the Stanford COVID-19 Biobank Study Group, Andra L Blomkalns, Samuel Yang

* Contributed equally

**Supplementary Methods**

|Q| dPCR cutoff for N1 and N2 –

Our positive call criteria were to have greater or equal to 2 positive partitions for both N1 and N2. Given that the |Q| array dPCR platform analyzes 95% of the 9µL input sample, detecting 2 copies in the 8·5µL analyzed results in 0·23 copies/µL.

|Q| limit of detection of viral RNA using |Q| Triplex Assay in Nasopharyngeal swab matrix–

We prepared 2x serial dilution of of AccuPlex™ SARS-CoV-2 Reference Material Kit from 1,600 copies/mL down to 50 copies/mL. 9 replicates of each dilution were assayed on the dPCR platform to determine the limit of detection at 200 copies/mL.

| Dilution of of AccuPlex™ SARS-CoV-2 Reference Material Kit | N1 positives ≥2 / total number of replicates | N2 positives ≥2 / total number of replicates |
| --- | --- | --- |
| 1600 cp/mL | 9/9 | 9/9 |
| 800 cp/mL | 9/9 | 9/9 |
| 400 cp/mL | 9/9 | 9/9 |
| 200 cp/mL | 9/9 | 9/9 |
| 100 cp/mL | 4/9 | 6/9 |
| 50 cp/mL | 2/9 | 4/9 |

|Q| limit of detection of viral RNA using |Q| Triplex Assay in plasma matrix–

We prepared 2x serial dilution of of AccuPlex™ SARS-CoV-2 Reference Material Kit from 800 copies/mL down to 0 copies/mL. 2 replicates of each dilution were assayed on the dPCR platform to determine the limit of detection. Our LOD would be 100 copies/mL were we to define a positive sample by either N1 or N2 ≥0·23. With our stringent cutoff of N1 and N2 ≥0·23, our LOD is 200 copies/mL.

| Dilution of of AccuPlex™ SARS-CoV-2 Reference Material Kit | N1 positives / total number of replicates | N2 positives / total number of replicates |
| --- | --- | --- |
| 800 cp/mL | 2/2 | 2/2 |
| 400 cp/mL | 2/2 | 2/2/ |
| 200 cp/mL | 2/2 | 2/2 |
| 100 cp/mL | 1/2 | 1/2 |
| 50 cp/mL | 0/2 | 1/2 |
| 0 cp/mL | 0/2 | 0/2 |

Using N1 or N2 <40 Ct for qPCR or ≥0·23 for dPCR on plasma would result in 36/147 (~24·5%) positive with qPCR and 68/91 (~35·6%) positive with dPCR.

Consistency and accuracy of the dPCR platform in detecting viral RNA –

We determined the consistency and accuracy of detection of viral RNA by the |Q| array dPCR platform by performing sets of serial dilutions of AccuPlex™ SARS-CoV-2 Reference Material Kit (SeraCare, CAT#0505-0126) in parallel. AccuPlex™ SARS-CoV-2 Reference Material Kit was diluted to 200, 400, and 800 copies/mL. 20 replicates of each dilution were run on the dPCR to determine that the consistency and accuracy of the platform was >95%.

**Supplementary Table**

**Table S1. Prediction of RNAaemia on presentation.**

|  | OR (95% CI) |
| --- | --- |
| **Sx: Cough** | **4·00 (1·39 - 14·58)** |
| Sx: Sore throat | 1·43 (0·55 - 3·56) |
| Sx: Chills | 1·23 (0·56 - 2·66) |
| Sx: Chest pain | 1·60 (0·75 - 3·42) |
| **ED: SpO2 low** | **2·80 (1·31 - 6·01)** |
| ED: Febrile | 2·04 (0·92 - 4·51) |
| N | 191 |
| AIC | 190·8 |
| AUROC | 0·66 |

Potential predictors of RNAaemia included: demographic features (age 60+ or 80+, sex), past medical history features (lung disease, cancer, diabetes, immunosuppression, heart disease, hypertension, angiotensin converting enzyme inhibitor or angiotensin receptor blocker use, stroke, dementia, deep venous thrombosis or pulmonary embolus, chronic kidney disease, tobacco smoking), binary indicators of abnormal ED vital signs (low or high mean arterial pressure, low or high heart rate, low or high respiratory rate, low oxygen saturation, low or high temperature), and patient-reported symptoms (fever, chills, cough, sore throat, congestion, shortness of breath, chest pain, myalgias, nausea/vomiting/diarrhea, loss of taste, loss of smell, confusion, headache).

To prevent over-fitting, variables were selected via elastic net regression of RNAaemia (1=detected, 0=not detected) on these features with 10-fold cross-validation, selecting the regularisation parameter λ minimizing mean cross-validated error, and yielding the features in the table above. In a logistic model regressing RNAaemia on these features, significant predictors of RNAaemia were: cough and low oxygen saturation by pulse oximetry (SpO2<95%). Mean cross-validated area under the receiver-operating characteristic curve (AUROC) of the model in predicting RNAaemia was 0·66.
